# Hospital Characteristics Associated with Observed Transcatheter Aortic Valve Replacement Prices

**DOI:** 10.1101/2024.05.20.24307421

**Authors:** Roni Ochakovski, Yuqi Zhang, Marcelo Cerullo

**Author notes:** **Funding:** Bass Connections, Duke University.

## Abstract

Transcatheter aortic valve replacement (TAVR) has emerged as a revolutionary treatment for aortic stenosis. However, TAVR prices vary considerably, and factors associated with this variation remain unclear. We aim to describe the variation in TAVR prices in relation to hospital financial performance among institutions ranked by the U.S. News and World Report (USNWR). Using a modified two-part model, we examined financial and operational characteristics (TAVR performance scores, median all-payer within-hospital TAVR price, net hospital profit margin, hospital markups [i.e., charge-to-cost ratio], bed days available, and CMS wage index) of 640 TAVR-performing hospitals ranked by the USNWR. After determining observed to expected (O:E) ratios for TAVR prices for each hospital, we then examined hospital characteristics across O:E quintiles. Overall, price disclosure was 48.6% (n=311). Between the lowest and highest O:E quintiles, median hospital markup (4.75 vs 5.33; p=0.41) and median net hospital margin (1.76 vs 3.15; p=0.12) were comparable. The highest O:E ratio quintile had lower median TAVR prices compared to the lowest O:E ratio quintile ($72,129.12 vs $49,022.03; p<0.001). Most significantly, TAVR price IQR’s within hospitals had a linear decline from the lowest to the highest O:E ratio quintiles ($119,043 vs $27,240; p<0.001). USNWR ranking scores had no significant variation across the quintiles (p=0.95). We concluded that hospitals that charge more than expected for TAVRs do not have higher profit margins nor markups and are not higher ranked by USNWR as those that charge less than expected. Additionally, with higher observed over expected TAVR prices, the variation in TAVR rates within hospitals decreased linearly. Finally, O:E TAVR price ratios appear to have no association with publicly reported hospital quality.

## Introduction

With the publication of findings from investigations in younger and lower-risk patients, transcatheter aortic valve replacement (TAVR) has begun to overtake surgical aortic valve replacement (SAVR) as the primary modality for the treatment of aortic stenosis. In addition to data supporting its long-term durability in lower risk patients, TAVR’s popularity has also grown considerably due to its lower procedural mortality rates and shorter hospitals stays (1). However, the up-front cost of TAVR is higher than that of SAVR, fueling a debate around the cost-effectiveness of each in relation to the other. While earlier literature justified TAVR’s use in selected intermediate to high-risk patients, more recent work has shown that the lower total costs of inpatient care and operating room resources offsets TAVR’s higher initial device and physician costs (2–3). Though negotiated payment rates for TAVR take into account these costs, TAVR prices vary widely across institutions (2). Some observers have argued that the higher price is driven by the higher quality of the hospitals which perform TAVRs (4–5). However, other commentators have noted that there is only a weak association between procedural pricing and hospital quality, and this phenomenon has been observed across myriad surgical procedures and therapies (6–12). Understanding the drivers of this variation in prices, as they may relate to procedural quality (if at all), is key to controlling the steep growth of healthcare costs and specifically for resource-intensive procedures such as TAVR.

To that end, the Centers for Medicare and Medicaid Services (CMS) has put into effect several regulations that allowed patients and payers to identify providers and facilities that have controlled outsized costs for procedures. Recently, a regulation termed the “Hospital Price Transparency Final Rule” that went into effect January 1, 2021, required hospitals to publish a machine-readable file accessible to patients (and researchers) that reports both self-pay and payer-negotiated prices for all procedures they perform (9). In spite of this, both the academic and lay press has documented extremely low compliance with these regulations (e.g., with some procedures having fewer than 25% of hospitals reporting prices) (9, 13). To our knowledge, hospitals’ compliance with this price transparency regulation has not yet been studied in the context of TAVR. TAVR is an ideal scenario to study resource-and technology-intensive hospital-based procedures for several reasons: (1) it is easily identifiable in claims and reimbursement codes, (2) it relatively “uniform” in its clinical implementation, and (3) has explicit requirements for Medicare reimbursement in terms of a facility’s surgical and clinical capabilities.

Our study seeks to explore the variation in TAVR prices as a function of hospital financial and operational characteristics using recently disclosed payment data in accordance with the CMS Price Transparency rule. To do so, we leverage several publicly available datasets containing pricing information with hospital quality metrics, while accounting for significant non-disclosure. We posit that the extent to which TAVR prices are disclosed is key to understanding overall variation in the prices and payments for TAVR, in part due to the complicated relationship between the “true cost” of a given procedure and the prices exacted for that procedure (14). We anticipate that our findings would buttress insights for crafting policy aimed at controlling and minimizing variation in procedural prices nationwide.

## Methods

Hospital procedural prices and respective disclosures were collected and organized by Turquoise Health, a public price transparency platform (15). Turquoise Health uses an algorithm to search hospital websites for publicly available pricing information. The prices disclosed are in line with 2021 Price Transparency Regulations (16).

The universe of TAVR-performing hospitals was provided by U.S. News and World Report (USNWR), which scores hospitals based on the quality of the procedure. The hospitals ranked by the USNWR were linked to their 2019 Medicare Cost Reports from the Healthcare Cost Report Information System (HCRIS) via Medicare Provider Number (17). The HCRIS data was used to provide the necessary financial health characteristics of the studied hospitals. The characteristics were analyzed for outliers and missing values were removed. The CMS wage index was also linked to this dataset via the Medicare Provider Number.

Within-hospital median TAVR price and IQR were calculated for each hospital using the Turquoise Data and the nine Current Procedural Terminology (CPT) codes and two Medicare Severity Diagnosis Related Groups (MS-DRG) codes. The CPT codes used were: 33361, 33362, 33363, 33364, 33365, 33366, 33367, 33368, and 33369. The MS-DRG codes were: 266 and 267 (Table 1). Then, a modified two-part model was applied in order to obtain a more universally comparable TAVR price adjusted for hospital characteristics (18–19). In the first part, the odds of price disclosure were regressed with descriptive characteristics of the hospitals (net hospital profit margin, hospital markups [i.e., charge-to-cost ratio], bed days available, government appropriations, total capital [fixtures and buildings], USNWR TAVR Quality Score, TAVR price median, and TAVR price IQR). Then this regression was transformed into a probability of price disclosure (as based on the above characteristics) and assigned individually to all TAVR hospitals. The second part of the model used this probability as well as the same hospital characteristics to calculate a predicted TAVR price for each hospital. From there, an expected TAVR price was assigned for each hospital. Finally, to form the adjusted TAVR prices, the observed TAVR price was divided by the expected TAVR price to create the observed to expected (O:E) ratio (*Appendix: Model Specification*). The O:E ratio was visualized and winsorized to the 5^th^ and 95^th^ percentiles.

**Table 1.**
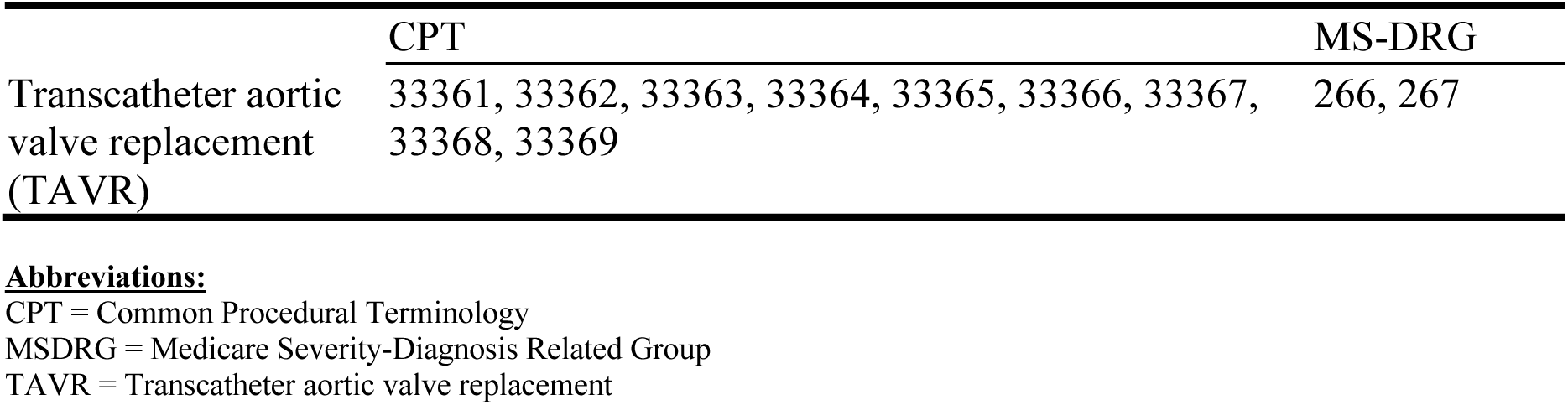
Common Procedural Terminology (CPT) and Medicare Severity-Diagnosis Related Group (MS-DRG) used to identify hospital prices for transcatheter aortic valve replacement (TAVR).

|  | CPT | MS-DRG |
| --- | --- | --- |
| Transcatheter aortic valve replacement (TAVR) | 33361, 33362, 33363, 33364, 33365, 33366, 33367, 33368, 33369 | 266, 267 |

**Abbreviations:**
CPT = Common Procedural Terminology
MSDRG = Medicare Severity-Diagnosis Related Group
TAVR = Transcatheter aortic valve replacement

Following this, the hospitals were organized into five quintiles based on their respective O:E ratios and eight hospital characteristics (USNWR TAVR performance scores, median all-payer within-hospital TAVR price, net hospital profit margin, hospital markups [i.e., charge-to-cost ratio], bed days available, and CMS wage index) were examined across these quintiles. The Wilcoxon-Sum Test was used to determine significant differences across quintiles. The within-hospital variation of TAVR prices (given by interquartile range [IQR]) was examined separately and also across O:E ratio quintiles; significance was measured using weighted least-squared regression. All analyses were performed using R Version 4.1.0 and at a significance level of p < 0.05. The study was approved by the Duke Institutional Review Board.

## Results

Exactly 640 hospitals were ranked by the USNWR in the TAVR-performing category. The median bed size was 397 (IQR=304). The median net patient revenue was $599,404,885 (IQR = $607,765,964). 490 (76.5%) of these were teaching hospitals. Additionally, 471 (76.1%) were non-profit, 61 (9.9%) were government owned and 86 (13.9%) were for-profit (Table 2).

**Table 2.**
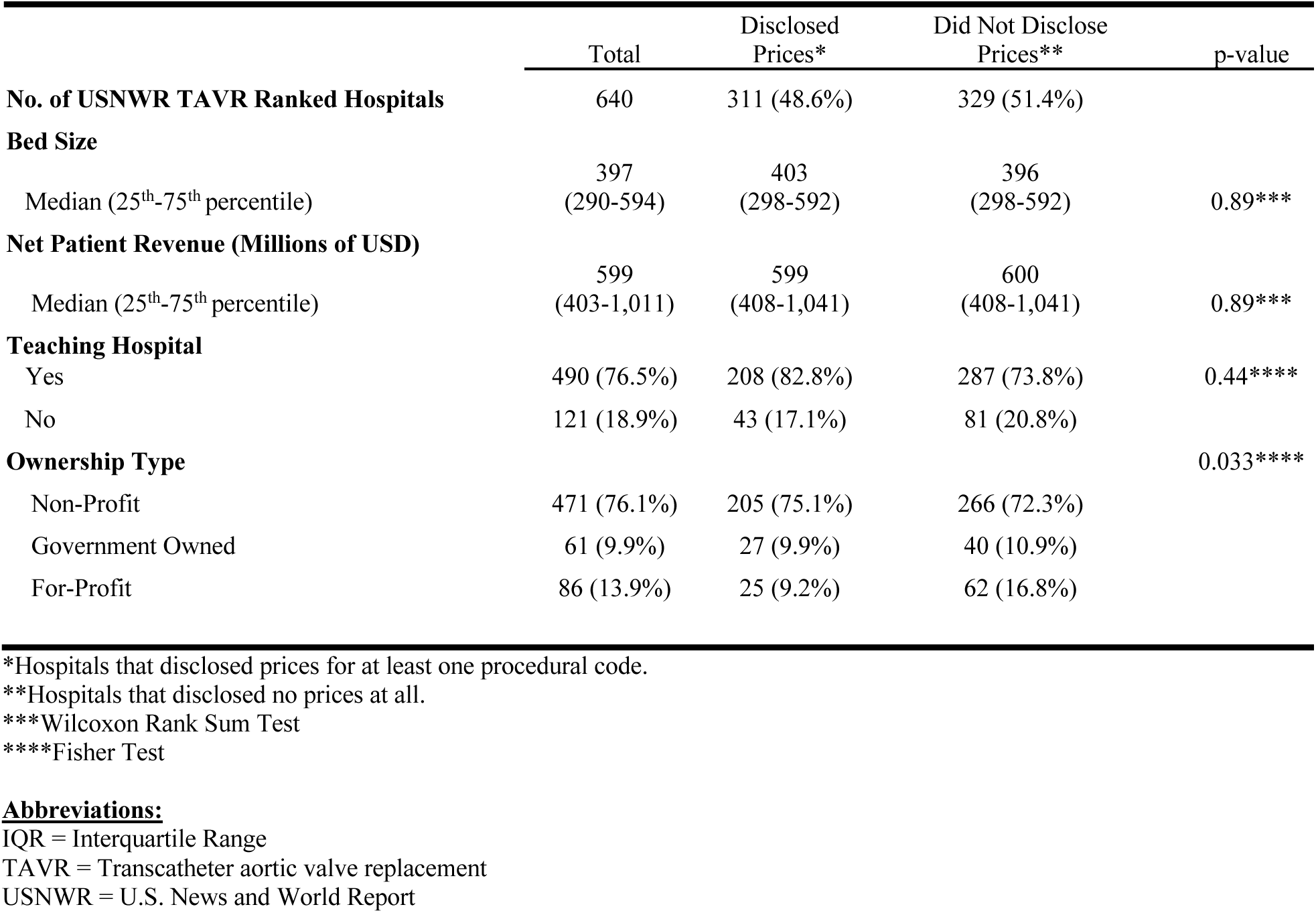
Characteristics of U.S. News and World Report (USNWR) TAVR Ranked Hospitals by Price Disclosures.

Of the USNWR-ranked hospitals, 311 (48.6%) disclosed prices for at least one of the procedures (p=0.89). There was no significant difference in median bed size between hospitals that disclosed and did not disclose prices (p=0.89). Disclosing hospitals had a median bed size of 403 (IQR = 270) and non-disclosing hospitals had 396 (IQR=317) median beds. The median net patient revenue of disclosing hospitals was $599,054,043 (IQR=$565,145,924) while that of non-disclosing hospitals was $600,606,110 (IQR=$631,704,912); the differences were insignificant (p=0.89). 208 (82.8%) of the disclosing hospitals and 287 (73.8%) of the non-disclosing hospitals were teaching hospitals (p=0.44). Differences in ownership were significant between disclosing and non-disclosing hospitals (p=0.033). For disclosing hospitals, 205 (75.1%) were non-profit, 27 (9.9%) were government-owned, and 25 (9.2%) were for-profit. For non-disclosing hospitals, 266 (72.3%) were non-profit, 40 (10.9%) were government-owned, and 62 (16.8%) were for-profit (Table 2).

In the first O:E quintile, there were 49 hospitals, 48 in the second, 50 in the third, 48 in the fourth, and 49 in the fifth. USNWR Score medians were mostly linear with the first quintile median being -0.148 (IQR=0.89) and the fifth quintile at -0.151 (IQR=0.87) (Fig 1). The median CMS wage index scores were also fairly linear across quintiles with the first quintile having an index of 1.00 (IQR=0.27), and 0.88 (IQR=0.13) in the last quintile (Fig 2). The net profit margin was $1.76 (IQR=$10.4) in the first quintile and $3.14 (IQR=$10.1) in the fifth quintile (Fig 3). The median markup in the first O:E quintile was 4.75 (IQR=2.4) and 5.33 (IQR=2.38) in the last quintile (Fig 4). The TAVR price median across O:E quintiles resembled a downwards parabola with the first quintile at $119,043 (IQR=$70,316), the third quintile at $69,361 (IQR=$29,031), and the fifth quintile at $28,240 (IQR=$19,699) (Fig 5). The median bed days available were also linear with the first O:E quintile at 163,155 (IQR=125,197) and the last O:E quintile at 27,240 (IQR=19,669) (Fig 6). The TAVR price IQR had a linear decline with the first quintile at $119,043 (IQR=$70,316) and the last quintile at $27,240 (IQR=$19,699) (Fig 7) (Table 3).

**Fig 1.**
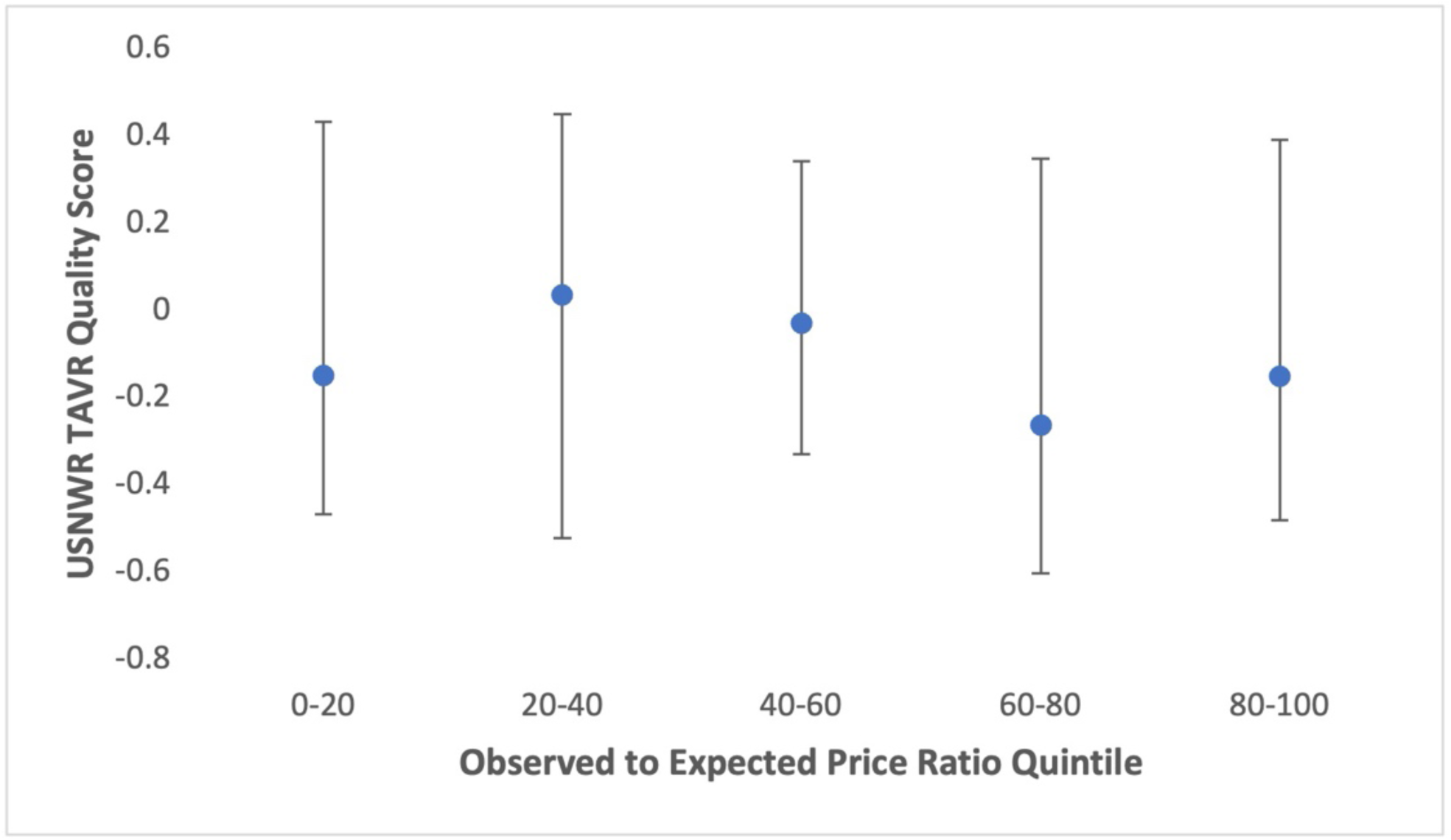
U.S. News and World Report Median TAVR Quality Scores Across Observed to Expected Transcatheter Aortic Valve Replacement Price Ratio **Legend:** Each point represents the median U.S. News and World Report quality score for transcatheter aortic valve replacement performing hospitals within each observed to expected price ratio. The error bars show the 25^th^ and 75^th^ percentiles of the quality scores.

**Fig 2.**
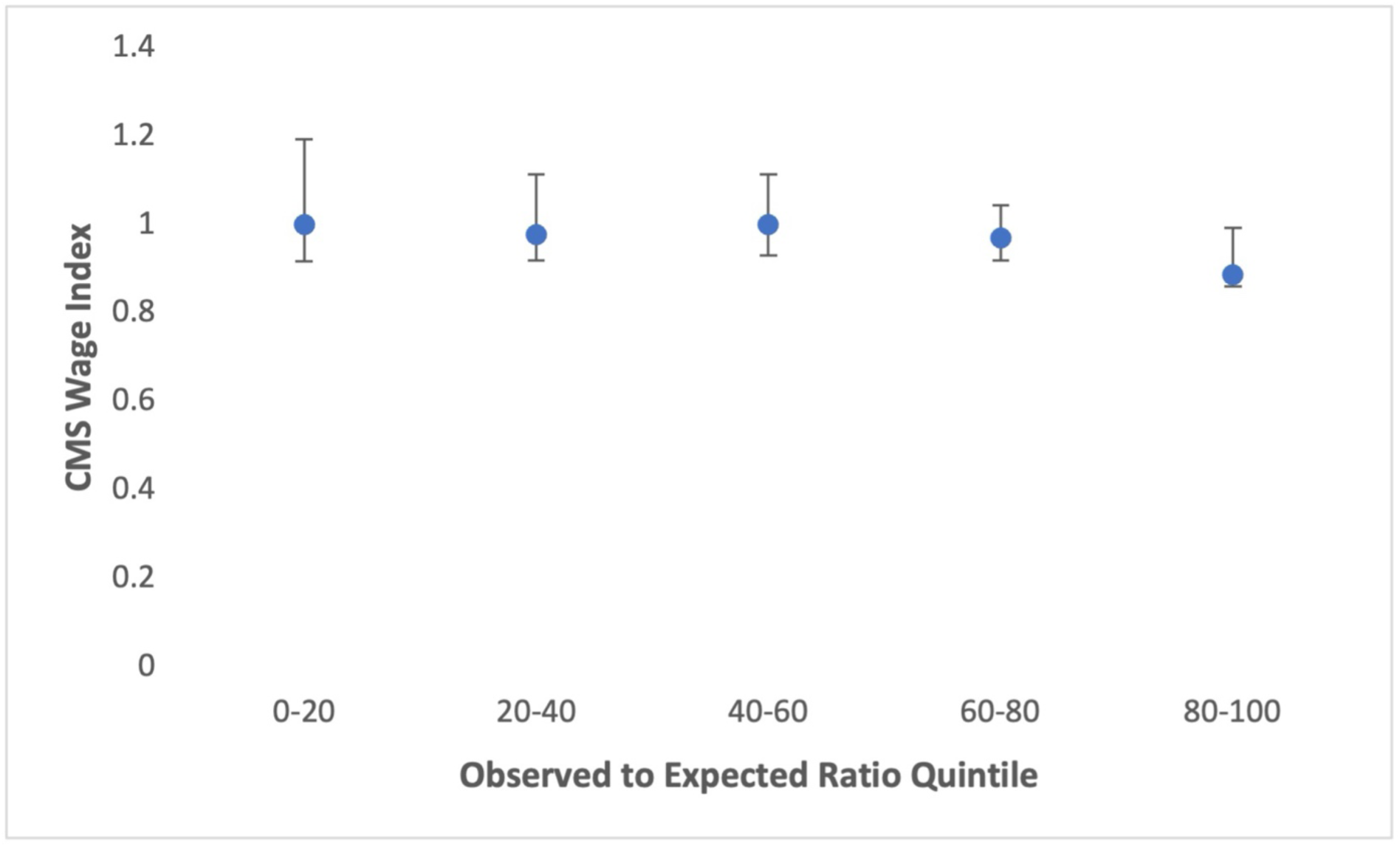
Hospital Net Profit Margin Across Observed to Expected Transcatheter Aortic Valve Replacement Price Ratio Quintiles **Legend:** Each point represents the median net profit margin of the hospitals in the respective observed to expected transcatheter aortic valve replacement price ratio quintile. The error bars show the 25^th^ and 75^th^ percentiles of the median profit margins.

**Fig 3.**
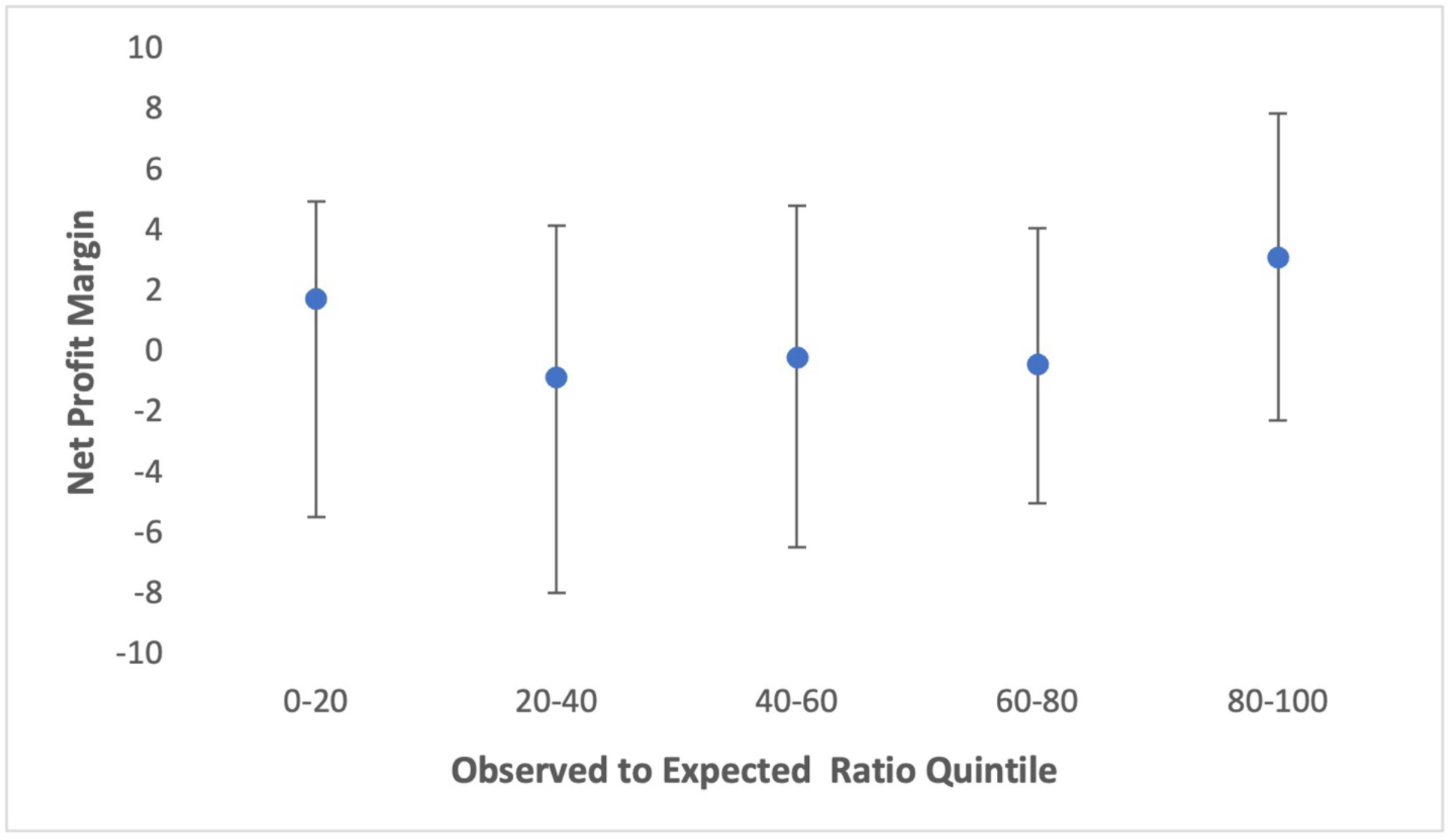
Median Markup Across Observed to Expected Transcatheter Aortic Valve Replacement Price Ratio Quintiles **Legend:** Each point represents the median markup of the hospitals in the respective observed to expected transcatheter aortic valve replacement price ratio quintile. The error bars show the 25^th^ and 75^th^ percentiles of the median markups. Markups were calculated as total hospital charges over total hospital costs.

**Fig 4.**
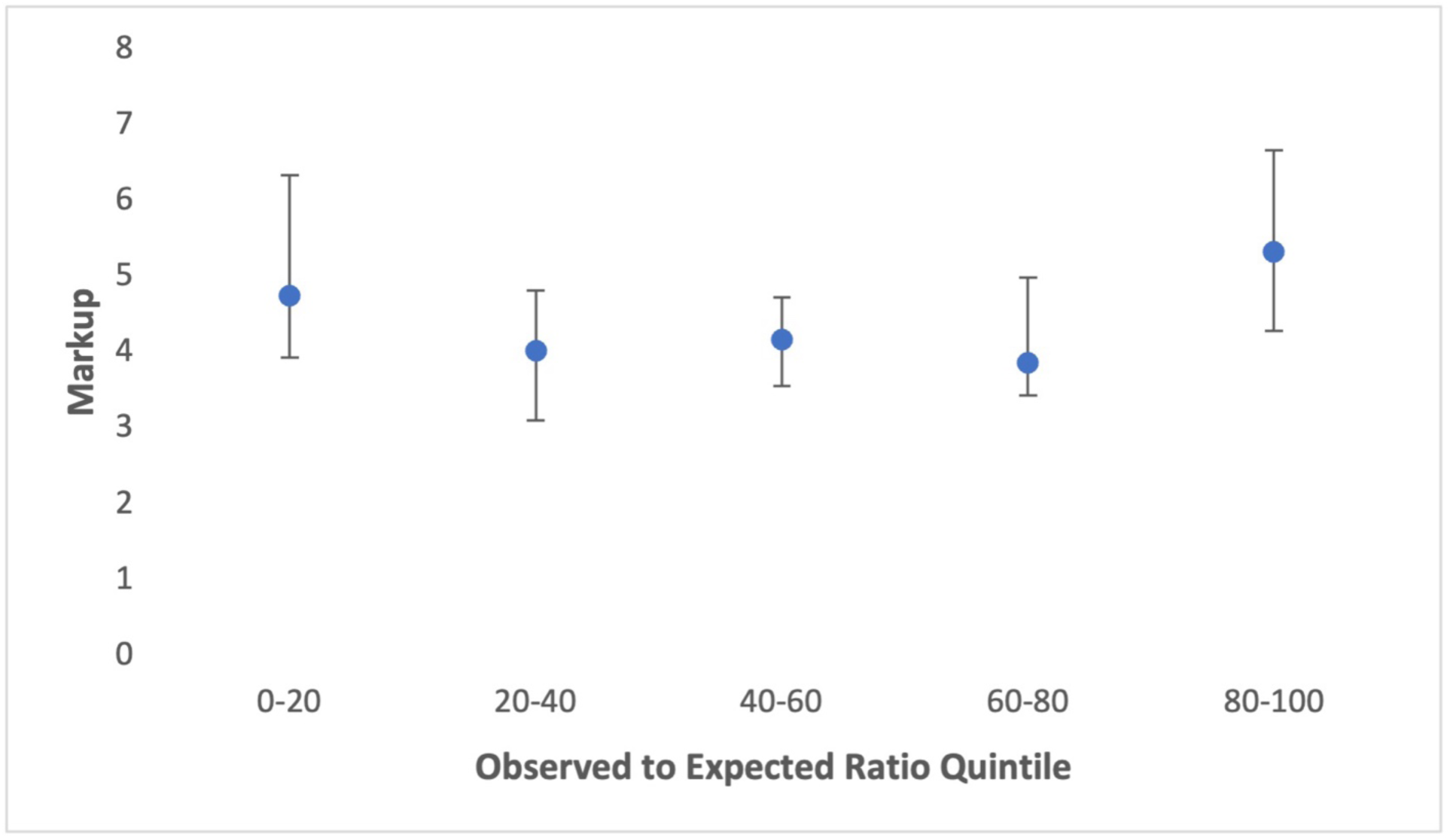
Median Bed Days Available Across Observed to Expected Transcatheter Aortic Valve Replacement Price Ratio Quintiles **Legend:** Each point represents the median bed days available of the hospitals in the respective observed to expected transcatheter aortic valve replacement price ratio quintile. The error bars show the 25^th^ and 75^th^ percentiles of the median bed days available. Bed days available were calculated as the number of beds available multiplied by the number of days in the calendar year.

**Fig 5.**
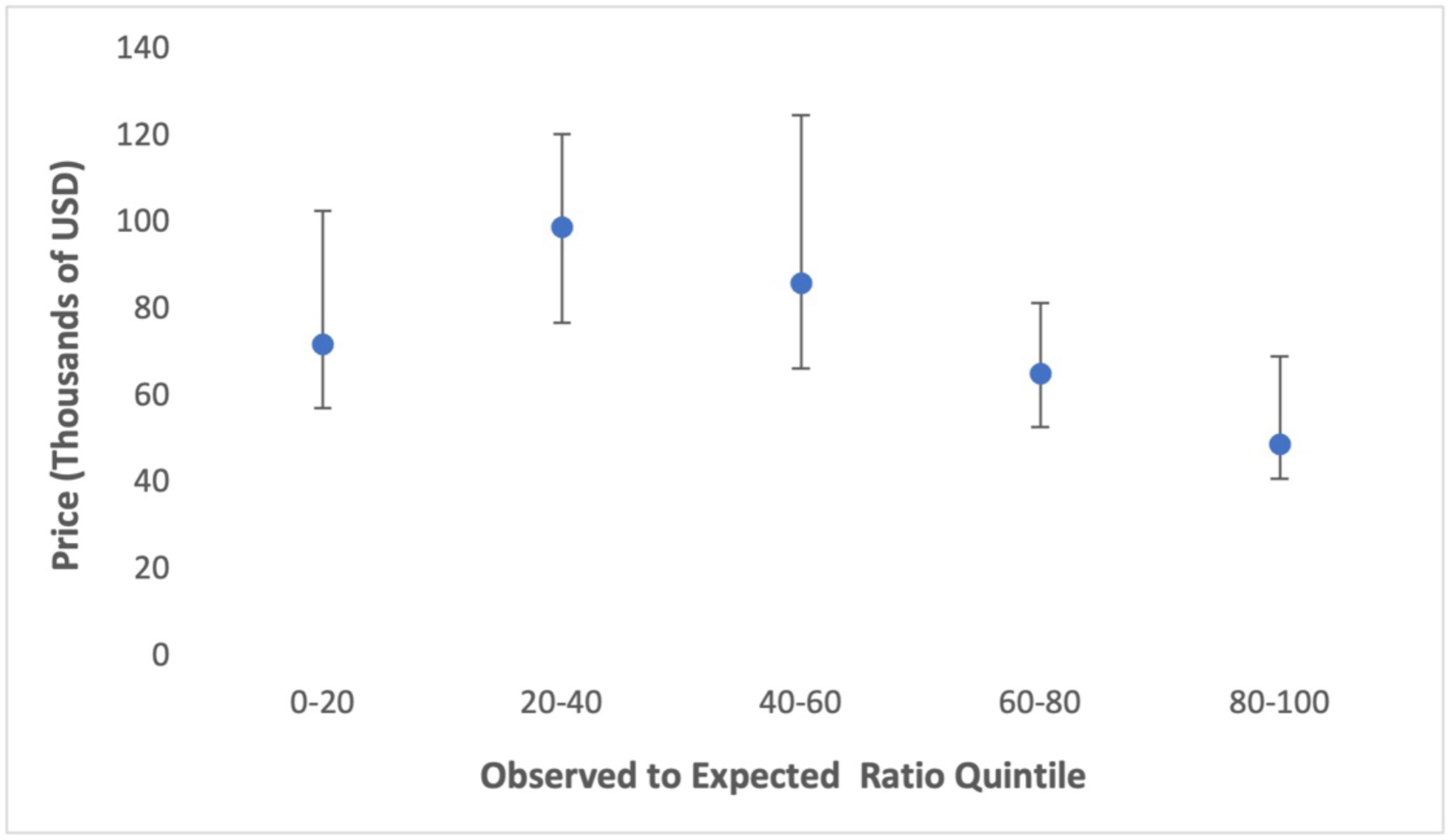
Median TAVR Price Across Observed to Expected Transcatheter Aortic Valve Replacement Price Ratio Quintiles **Legend:** Each point represents the across-payer median transcatheter aortic valve replacement price of the hospitals in the respective observed to expected transcatheter aortic valve replacement price ratio quintile. The error bars show the 25^th^ and 75^th^ percentiles of the median prices.

**Fig 6.**
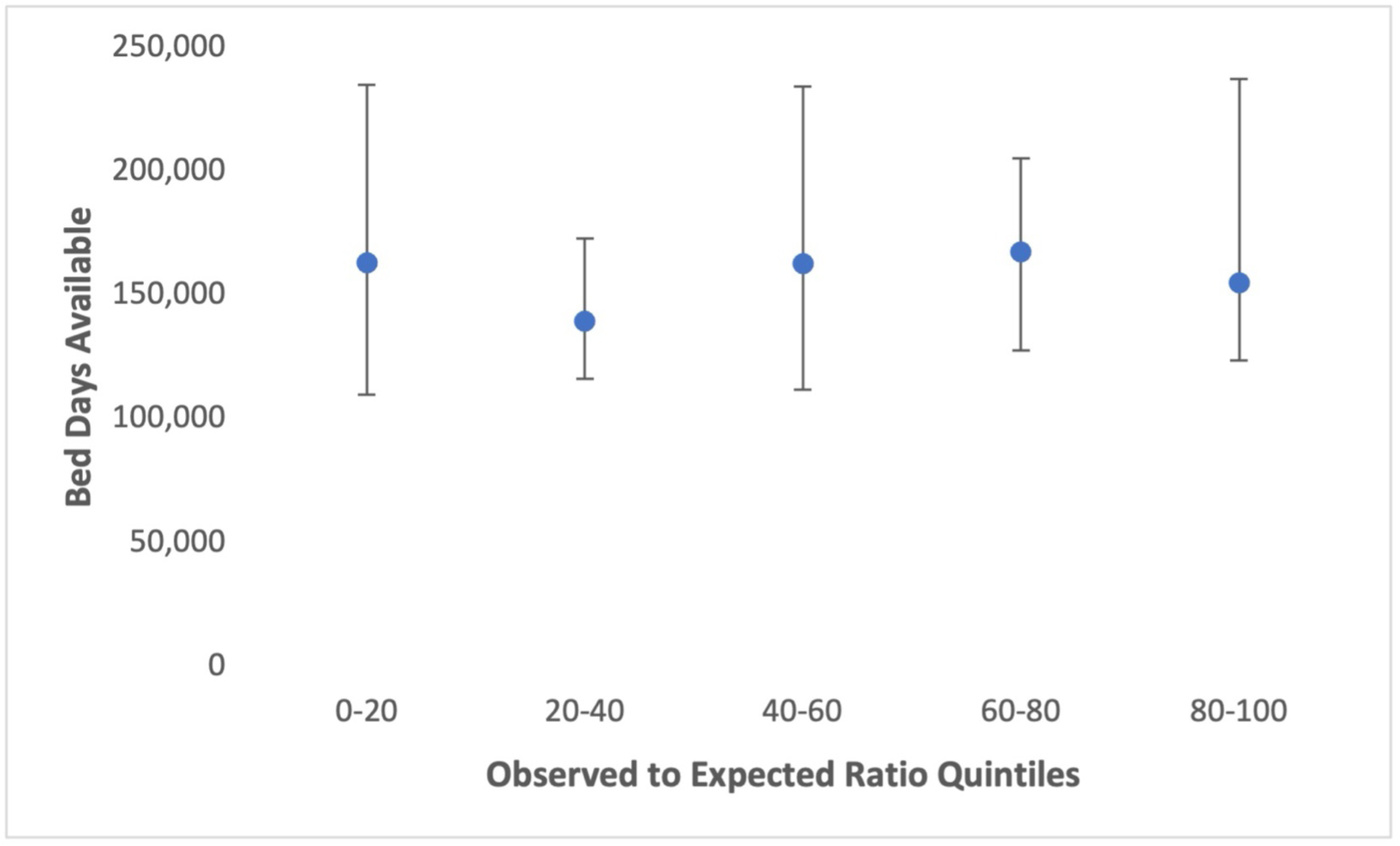
Median US Centers for Medicare & Medicaid Services Wage Index Across Observed to Expected Transcatheter Aortic Valve Replacement Price Ratio Quintiles **Legend:** Each point represents the median US Centers for Medicare & Medicaid Services Wage Index for hospital regions in the respective observed to expected transcatheter aortic valve replacement price ratio quintile. The error bars show the 25^th^ and 75^th^ percentiles of the median wage indices.

**Fig 7.**
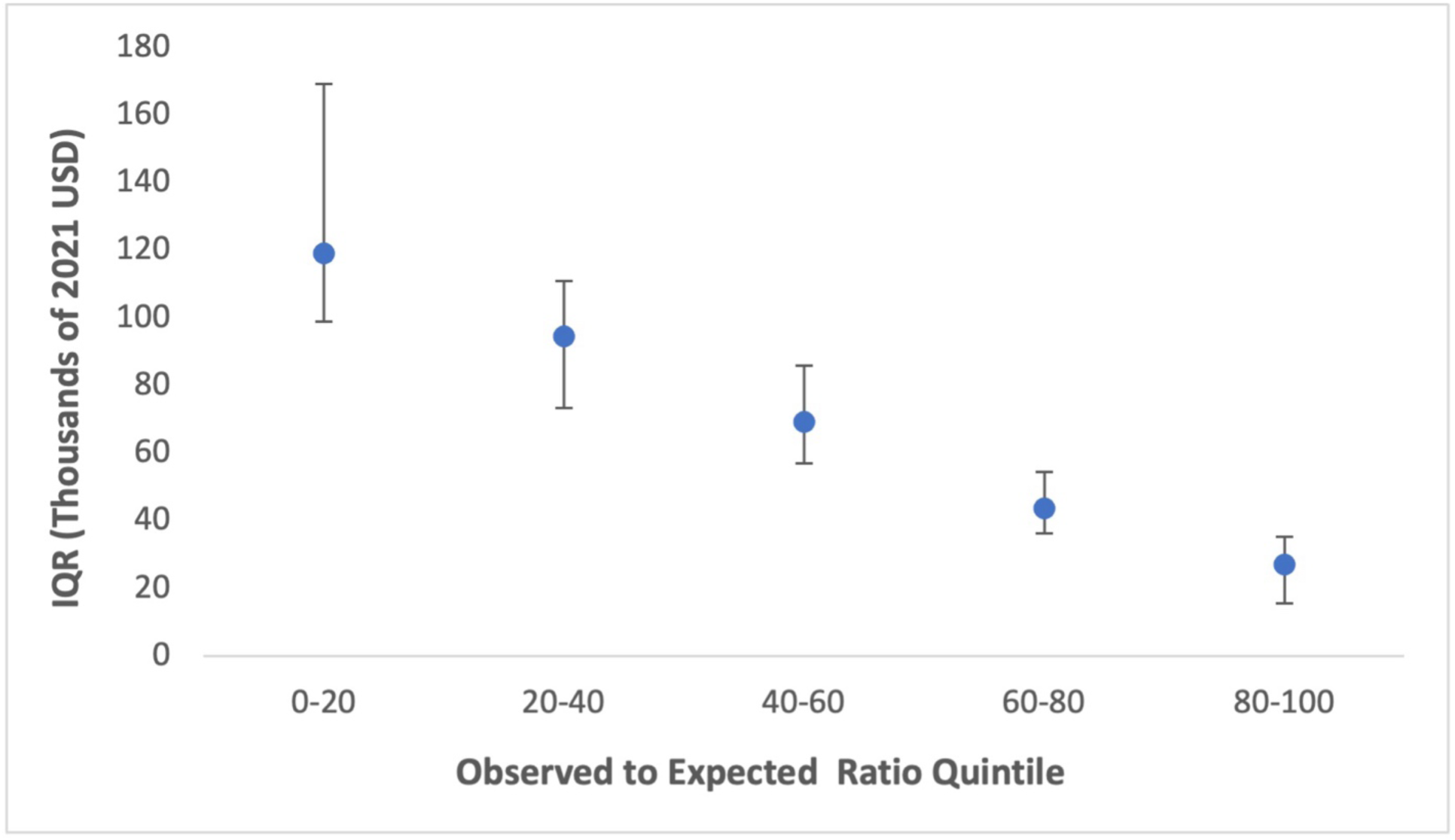
Hospital Interquartile Range Across Observed to Expected Transcatheter Aortic Valve Replacement Price Ratio Quintiles **Legend:** Each point represents the median interquartile range of the hospitals in the respective observed to expected transcatheter aortic valve replacement price ratio quintile. The error bars show the 25^th^ and 75^th^ percentiles of the median interquartile range. The points were calculated as the median of the interquartile ranges of prices (across payers) within a hospital.

**Table 3.** Hospital Characteristics Across O:E Ratio Qutiles.

|  | Median (IQR) |  |  |  |  | p-value** |
| --- | --- | --- | --- | --- | --- | --- |
|  | Quintile 1 | Quintile 2 | Quintile 3 | Quintile 4 | Quintile 5 |  |
| USNWR* TAVR Performance Score | -0.15 (0.89) | 0.036 (0.97) | -0.029(0.67) | -0.26 (0.95) | -0.15 (0.87) | 0.95 |
| Median TAVR Price | 72,129.12 (45,552) | 99,005.82 (43,611) | 86,177.5175 (58,452) | 65,358.02 (28,521) | 49,022.03 (28,163) | <0.001 |
| Net Profit Margin | 1.76 (10.4) | -0.82 (12.1) | -0.16 (11.3) | -0.39 (9.1) | 3.14 (10.1) | 0.12 |
| Markups | 4.75 (2.4) | 4.02 (1.7) | 4.17 (1.17) | 3.86 (1.56) | 5.33 (2.38) | 0.41 |
| Bed Days Available | 163,155 (125,197) | 139,430 (56,540) | 162,790 (122,574) | 167,541 (77,656) | 154,967 (113,837) | 0.8 |
| CMS Wage Index | 1.00 (0.27) | 0.97 (0.19) | 1.00 (0.18) | 0.97 (0.12) | 0.88 (0.13) | 0.002 |
| TAVR Price IQR | 119,043 (70,316) | 94,668 (37,762) | 69,361 (29,031) | 43,708 (18,034) | 27,240 (19,669) | <0.001*** |

\*U.S. News and World Report
\*\*Wilcoxon Rank Sum Test
\*\*\*Weighted Linear Regression

The USNWR scores had no significant relationship across O:E quintiles (p=0.95) (Fig 1). This was also reflected in the net profit margin (p=0.12) (Fig 3), markups (p=0.41) (Fig 4), and bed days available (p=0.80) (Fig 6). However, there was a significant difference between the highest and lowest quintiles for median TAVR prices (p<0.001) (Fig 5) and CMS wage index (p=0.002) (Fig 2). The TAVR price IQR also had a significant relationship with O:E quintiles (p<0.001). As O:E quintile increased, TAVR price IQR decreased by $8,548 (95% CI: -112,15.49 to -$5,880.54) on average (Fig 7) (Table 3).

## Discussion

This study had four principal findings that improve the understanding of the relationship between price transparency regulations and hospital financial characteristics of TAVR-performing hospitals. First, we found that hospitals that charge more for TAVRs do not have higher profit margins nor markups and are not higher ranked by USNWR as those that charge less. Second, with higher TAVR prices, the variation of within-hospital TAVR prices decreased linearly. Third, O:E TAVR price ratios appear to have no association with publicly reported hospital quality. And fourth, hospitals that charged higher TAVR prices were more likely to be located in regions with lower incomes.

Our first finding indicates that hospital TAVR prices have no significant relationship with hospital profit margins or markups. Therefore, hospitals don’t appear to directly benefit from charging higher prices. This stands in contradiction to prior work which has noted both a strong association between profit margins and markups as well as between costs of procedures and net income (20–21). One study looked at Medicare Cost Reports for 2,993 acute care hospital and found that hospitals with higher markups are statistically more likely to be more profitable than those with lower markups (20). In another retrospective analysis of hospital profitability, researchers found that of the 2,824 hospitals studied, a significant portion have relied on increased prices in order to drive higher profits, rather than decrease costs (21). However, in contrast to the all-encompassing analysis of hospital revenues, our study focused on a prime example of an expensive, variable, yet shoppable procedure (TAVR). This allowed us to address the limitations of prior studies regarding the differing effects of shoppable versus non-shoppable procedures on revenues. Additionally, by using O:E ratios rather than absolute prices, we were able to provide a novel approach to universally weighing prices by a whole subset of hospital characteristics (*Appendix: Model Specification*).

Second, we noted as hospitals charge more for TAVRs, the variation of TAVR prices decreases. We propose two possible explanations for this. First, the variation in prices could reflect real price differences as influenced by factors we do not account for in our model.

However, given the comprehensive nature of our model, the second explanation seems more likely. We hypothesize that there must be a maximum TAVR price, beyond which it becomes unreasonable to charge payers. Ergo, as hospitals charge more and approach said maximum, the variation in prices decreases. Such a phenomenon would also explain the linear nature of the variation decline as prices increases. Similar to our study, many others have noted significant variation across hospitals when looking within the same procedure. In one study, researchers looked at a variety of common shoppable procedures such as Magnetic Resonance Imaging, cardiac surgery, and joint replacements and found up to a 10-fold difference in prices in procedures across all price-disclosing hospitals (10). Another study explored prices of common shoppable surgical procedures and had similar findings that along with low compliance rates, some procedures reached interquartile ranges of up to 1.7 times the median (9). Our study extends these previous findings by attempting to explain this pattern of variation as seen in TAVRs.

Third, we found there to be no association between TAVR prices charged and the hospital quality as reported by the USNWR. This implies that patients may have more choices in shoppable procedures such as TAVRs as quality is not associated with cost price. This mirrors previous literature, which also found no association between quality of hospitals and the prices charged^22^. More specifically, one study exploring coronary artery bypass graft procedures found no association between the quality of the hospital performing the procedure and the price of the procedure (6). Another source found that higher costs in hospitals were actually associated with higher rates of complications in procedures, leading to the same conclusion as before (7).

Finally, we found that higher-charging hospitals were located in areas with lower wages than those that charged lower prices. Although literature connecting hospital prices and demographics of patients is sparse, we offer several explanations. First, insurance market concentration is most likely lower in low-income areas giving hospitals more market power.

Second, hospitals may incur more uncompensated costs in lower-wage areas and may have higher procedure prices to balance out revenues. Third, the unique power dynamics between hospitals and low-income patients may provide the opportunity for hospitals to charge more than they otherwise would.

Our study should be interpreted in the context of several limitations. First, our data was gathered by Turquoise Health which mined all publicly available machine-readable hospital price data. However, as with most data mining protocols, random error could have contributed to missing or misinterpreted data points. The dataset could have been missing hospitals that did not report prices or reported prices incorrectly. Additionally, some price data was reported by hospitals as percentages of a gross cash price but collected as a decimal smaller than one. To mitigate this bias, all prices smaller than $100 were removed from the data prior to analysis.

Furthermore, given the results and patterns were derived from a TAVR price dataset, conclusions are limited in their applicability to other procedures. However, prior evidence points to most shoppable surgical procedures having common financial attributes, thereby expanding the application of our results (7, 9, 10). Furthermore, for the CMS Wage Index and TAVR median prices, we compared the first and last quintiles to establish a significant association. Even so, there was significant “noise” in the middle quintiles such that a linear relationship could not be fully established. Nevertheless, this does not negate differences between the most extreme quintiles. Finally, we used the USNWR-ranked hospitals as our universe of TAVR-performing hospitals, which may have excluded some lesser-known hospitals from our analysis. Nonetheless, given the exploratory nature of the study, it is unlikely that more price data from other TAVR-performing hospitals would have had significant effects on our results.

This study has major implications for patients and policy makers. First, in line with previous studies, compliance with price transparency regulation remains low. This calls for stricter regulations and harsher fines for non-compliant hospitals. Such regulations would have a significant effect especially for shoppable services such as TAVRs by better informing patient choice and providing higher-value services. Second, in our study we have shown that regulation that would aim to stabilize TAVR prices would have minimal effects on hospital profits as hospitals with the highest variation had the lowest observed to expected prices. Additionally, given there was no association between observed to expected prices and quality, it shows that even for extremely costly procedures, patients may be better off comparing their options solely based on price and convenience rather than “quality”. Finally, provided there was no significant relationship between hospital profit margin and observed to expected prices, further price-stabilizing regulations or price-ceilings would be expected to have minimal effects on hospitals’ bottom lines while decreasing variation in prices and improving accessibility of shoppable surgical procedures for low-income areas.

## Supporting information

Supplemental Appendix 1 Model Specification

## Data Availability

All data produced in the present study are available upon reasonable request to the authors.

## ACKNOWELDGEMENTS

We would like to thank Paul R. Sabharwal and Samantha Owusu-Antwi for their analytic insights, Duke Bass Connections for their generous support, and Duke Undergraduate Research Support Grant for facilitating conference attendance.

