## Supplemental Appendix 1 Model Specification for "Hospital Characteristics Associated with Observed Transcatheter Aortic Valve Replacement Prices"

**S1 Appendix. Model Specification**

The first part of the model involved creating a binomial model predicting the odds of TAVR price-disclosure as follows:

$$TAVR\ Price\ Disclosure_i = \beta_0 + \beta_1 USNWR\ TAVR\ Quality\ Score_i + \beta_2 Hospital\ Profit\ Margin_i + \beta_3 Markup_i + \beta_4 Bed\ Days\ Available_i + \beta_5 TAVR\ Rate\ IQR_i + \beta_6 Government\ Apporpriations_i + \beta_7 Total\ Capital\ (Fixtures)_i + \varepsilon_i$$

$$i = individual\ hospital$$

Then, using the results from the first part of the model, we extracted a probability of TAVR price disclosure, $\hat{p}$. Then for the second part of our model, we used a Log-Linked Gamma Model to model the median TAVR price only within the hospitals that disclosed prices:

$$Median\ TAVR\ Price_i = \beta_0 + \beta_1 USNWR\ TAVR\ Quality\ Score_i + \beta_2 Hospital\ Profit\ Margin_i + \beta_3 Markup_i + \beta_4 Bed\ Days\ Available_i + \beta_5 TAVR\ Rate\ IQR_i + \beta_6 Government\ Apporpriations_i + \beta_7 Total\ Capital\ (Fixtures)_i + \varepsilon_i$$

$$i = individual\ hospital$$

Following that, we calculated each hospital's expected TAVR price:

$$Predicted\ TAVR\ Price_i = \hat{p}_i \times Median\ TAVR\ Price_i$$

$$i = individual\ hospital$$

Finally, the observed to expected TAVR price ratio was calculated:

$$Observed\ to\ Expected\ TAVR\ Price\ Ratio_i = \frac{Actual\ TAVR\ Price_i}{Predicted\ TAVR\ Price_i}$$

$$i = individual\ hospital$$
